# The emergence of superficial dermatophytosis due to *Trichophyton indotineae* and *Trichophyton mentagrophytes* genotypes VII and II* in the United States: A need for comprehensive testing approaches

**DOI:** 10.1101/2025.11.14.25340007

**Authors:** Gabrielle Todd, Vinay Vaida, Brittany O’Brien, Sudha Chaturvedi

## Abstract

We report an exponential rise in dermatophyte infections belonging to the *Trichophyton interdigitale/mentagrophytes* species complex (*TiTm*SC), including the recently speciated *T. indotineae* (*Ti*, formerly *T. mentagrophytes* genotype VIII), in New York City and surrounding counties. These include 135 cases of *Ti*, 39 cases of *T. mentagrophytes* genotype VII (*Tm*VII), and 14 cases of *T. mentagrophytes* genotype II* (*Tm*II*), highlighting the urgent need for rapid and high-throughput identification, and characterization of the prevalence, spread, and drug-resistance of these pathogens. A deep learning model utilizing a Convolutional Neural Network (CNN) was developed with the internal transcribed spacer (ITS) region of 28 genotypes of *TiTm*SC. The model enabled accurate, rapid, and high-throughput identification of *Trichophyton* species and genotypes directly from Sanger sequencing output files. Antifungal susceptibility testing (AFST) of these pathogens revealed that 57% of the *Ti* isolates were resistant to the first-line drug terbinafine, and 38-52% of *Ti* isolates were resistant to azoles. In contrast, all *Tm*VII and *Tm*II* isolates were susceptible to terbinafine, with some degree of resistance observed against azoles. Whole-genome sequencing (WGS) on a subset of *Ti*, *Tm*VII, and *Tm*II* isolates revealed both clonal spread and independent introductions. The comprehensive approaches on rapid identification, drug resistance profiles, and mechanisms of spread of these emerging dermatophyte pathogens will likely aid in patient care and infection control measures.

**IMPORTANCE:** In this report, we present a comprehensive laboratory investigations on the emergence and rapid spread of drug-resistant *Trichophyton indotineae* (*Ti*), *T. mentagrophytes* genotype VII (*Tm*VII) and *T. mentagrophytes* genotype II* (*Tm*II*) in New York City and surrounding counties. The whole-genome sequencing results revealed both clonal spread and independent introduction. The comprehensive approaches on rapid identification, drug-resistance profiles, and mechanisms of spread of these emerging pathogens will likely aid in patient care and infection contrl measures.

## INTRODUCTION

Dermatophytosis is a highly prevalent disease that impacts 20-25% of the population worldwide and, in many parts of the globe, infections are currently being driven by members of the *Trichophyton interdigitale/mentagrophytes* species complex (*TiTm*SC) (1–3). Taxonomic classification of these pathogens has shifted over time; *T. interdigitale* is considered anthropophilic, induces low levels of inflammation and is the main cause of tinea uguium and tinea pedis, while *T. mentagrophytes* is generally considered zoophilic or geophilic and generates highly inflammatory lesions on other parts of the body (1, 4–6). To date there are 28 known genotypes of the *TiTm*SC, which cannot be differentiated solely based on phenotypic characteristics and must be distinguished by one to nine single nucleotide polymorphisms (SNPs) in the internal transcribed spacer (ITS) region (1, 5, 6). Different members of the *TiTm*SC infect different body parts, have different mechanisms of spread, affect populations in different parts of the world and have different drug resistance profiles (6). Thus, genotyping suspected *TiTm*SC isolates is critical to identify the infection source, monitor spread and inform clinical treatment options (1).

*T. mentagrophytes* genotype VIII was ultimately given its own species name, *T. indotineae* (*Ti*), due to its high resistance to the first-line drug terbinafine and its rapid spread throughout India (7). Although this new species was previously categorized as *T. mentagrophytes* and a few cases have been detected in animals (8, 9), it appears to easily spread from person to person and there is one known instance of likely sexual transmission (10–12). Since its speciation in 2018, *Ti* has spread globally (8, 13–19). *Ti* causes extensive, pruritic plaques, typically on the trunk, extremities, and groin that are frequently resistant to the first-line drug, terbinafine (15, 18, 20). Infections may require prolonged treatment (e.g., ≥2 months) with second-line drugs such as itraconazole or other antifungals typically reserved for invasive fungal infections (21, 22). Patients of all ages and genders are affected by *Ti* infection. It is hypothesized that the emergence and spread of *Ti* is being driven by inappropriate use and overuse of over-the-counter topical creams containing combinations of antifungals, antibacterials, and high-potency corticosteroids (8, 23, 24).

The *Trichophyton mentagrophyte genotype* VII (*Tm*VII) infections, which manifest as pruritic, annular, scaly lesions on areas such as the face, buttocks, and genitals, are mainly diagnosed in men who have sex with men (MSM) (25). These lesions can be particularly severe, causing discomfort and negatively impacting the quality of life for the affected individuals. *Tm*VII infection was first reported in MSM in France and previously in men who traveled to Southeast Asia for sex tourism (25, 26). *Tm*VII is believed to spread through sexual contact and often requires prolonged antifungal treatment. No drug resistance has yet been observed in *Tm*VII to the first-line (terbinafine) or second-line (itraconazole) (27).

While *Trichophyton mentagrophyte* genotype II* (*Tm*II*) is considered as a less urgent pathogen than *Ti* or *Tm*VII, it is still a cause for concern due to its ability to cause inflammatory lesions like tinea capitis and tinea corporis (1). Importantly, as of now, no resistance against terbinafine or other antifungal drugs has been reported in *Tm*II*, providing a strong sense of security about treatment options (1).

We have previously reported the emergence of *Ti* and *Tm*VII in New York City (NYC), United States (US). The first two cases of *Ti* were reported in May 2023, followed by a cohort of 11 cases reported in June 2024 (14, 15). Similarly, the first case of *Tm*VII was reported in 2024, followed by four cases in the same year (27, 28). This emergence is significant as it marks the first documented cases of these pathogens in the US, highlighting the need for increased awareness and understanding of the diseases they cause. To the best of our knowledge, infections with *Tm*II* have thus far not been reported in the US.

Traditional culturing methods are unable to identify *Trichophyton* pathogens at the species and genotype levels, which presents a significant challenge for low-complexity clinical laboratories. Given the large number of genotypes (at least 28) within the *TiTm*SC, sequencing of the ITS region is the ideal method for species and genotype identification (29). Deep learning tools have shown great promise in identifying the infectious agents, including the dermatophytes (30, 31), underscoring the urgency for adopting new techniques in pathogen identification.

In this study, we have employed comprehensive laboratory approaches, including a deep learning model for rapid and high-throughput *Trichophyton* species and genotype identification directly from Sanger sequencing files, AFST with a broad range of antifungal drugs to determine resistance profiles, and WGS to determine the relatedness and spread of *Ti*, *Tm*VII, and *Tm*II* in the human population. We chose to focus on these three *Trichophyton* species and genotypes because their incidence in the greater NYC area has rapidly increased over the past three years. The data provided in this study enhances our understanding of these pathogens and will aid in patient care and infection control measures.

## MATERIALS AND METHODS

### Patient data

This investigation includes 188 unique patients infected with *Ti* or *Tm*VII or *Tm*II*. All patients came from the greater NYC metropolitan area. Patient demography included age, sex, and county of residence. Only dermatophytes *Ti*, *Tm*VII and *Tm*II* from the *TiTm*SC were included in this investigation. This study encompassed public health surveillance activities and is exempt according to New York State Department of Health Institutional Review Board guidelines.Informed consent was waived because the data were deidentified.

### Isolate culture and identification

Suspected dermatophyte isolates received from various healthcare facilities in NYC and surrounding counties were cultured on Sabouraud dextrose agar for three to 10 days at 30°C, and on potato dextrose agar at 30°C for seven to 10 days to induce sporulation.

Isolate identification to species and genotype levels was done by DNA sequencing of the ITS region of the ribosomal gene (15). Genomic DNA from each suspected *Trichophyton* isolate was extracted using a QIAamp DNA Mini Kit (Qiagen Inc., Cat #51306). A sterile needle was used to transfer culture to a tube prefilled with beads and lysis buler. Following a one-hour incubation at 70°C in a thermomixer (Eppendorf, Thermomixer C Model 5382) at 1000 rpm, tubes were placed in a Precellys homogenizer (Bertin Technologies, Cat #P002511-PEVT0-A.0) at 6500 rpm 3×60 s with a 15 s interval. The lysate was centrifuged briefly to reduce foaming, transferred to a new tube and DNA was extracted on QIAcube automated DNA extractor (Qiagen Inc., Cat #9001292). Subsequently, the ITS region was amplified using the primer set V1827 (5’-GGAAGTAAAAGTCGTAACAAGG - 3’) and V50 (5’ – TCCTCCGCTTATTGATATGC - 3’) using AccuTaq ™ LA DNA Polymerase (Sigma, Cat #D5553) and Sanger sequenced by the Wadsworth Center Advanced Genomic Technologies Cluster. For manual analysis of ITS sequences, forward and reverse sequences were trimmed and aligned using Sequencher (Gene Codes Corporation, Ann Arbor, MI, US) and a BLAST search of the consensus sequence was performed at the National Center for Biotechnology Institute (https://blast.ncbi.nlm.nih.gov/Blast.cgi) and at the Westerdijk Fungal Biodiversity Institute (https://wi.knaw.nl/page/Pairwise_alignment) to identify the isolate to the species level. Identification down to the genotype level was done by aligning ITS sequences to all 28 known genotypes in the *TiTm*SC (including *Ti*) (5) using CLC Genomics Workbench (version 25.0.3; Qiagen, Inc.). To accurately speciate and assign a genotype, the entire length of the ITS1-5.8S-ITS2 region comprising all unique *TiTm*SC SNPs (593-596 base pairs depending on the genotype) was used. Importantly, the very first nucleotide of the ITS1 region is essential for speciation: there is a “G” at this location for all *T. interdigitale* genotypes and an “A” for all *T. mentagrophytes* genotypes (Supplementary Figure 1). Only isolates with a 100% match to one of the 28 known *TiTm*SC ITS sequences were included in further analyses.

### Antifungal susceptibility testing

Antifungal susceptibility testing (AFST) was performed by a broth microdilution assay according to Reference Method M27-A3 of the Clinical and Laboratory Standards Institute (CLSI). Minimum inhibitory concentrations (MICs) were determined after 96 hours of incubation at 35°C as reported previously (15). Resistance against certain antifungals is based on available epidemiological cutol values (ECV) (17, 32, 33).

### Automated pipeline for *Trichophyton* species and genotype determination

An ITS-based deep learning model for rapid and accurate *Trichophyton* species and genotype identification was developed using 28 curated reference ITS sequences from the *TiTm*SC containing *Ti,* six genotypes of *T. interdigitale,* and 21 genotypes of *T. mentagrophytes* (5). The detailed steps required for feature engineering, training and prediction of the model are provided in Figure 1. In brief, for feature engineering, we computed a 6-mer count of tokens for each of the 28 *TiTm*SC ITS sequences. The choice of 6-mers was based on previous findings that 6-mers are the minimum length that captures informative signals for fungal ITS classification (34). These 6-mer counts were then normalized using standard z-score scaling to ensure that no k-mer frequency dominated and to ensure stable, elicient, and balanced learning across features, making the training process faster and more reliable. For making Convolution Neural Network (CNN) model architecture, an input layer was followed by two convolutional layers with Rectifier Linear Unit (ReLU) activation, a flattening layer, a fully connected dense layer with ReLU activation, and a final output layer with SoftMax activation across 28 *TiTm*SC ITS sequences. For model hyperparameters, we used categorical cross-entropy loss to optimize 28 *TiTm*SC ITS sequences. Adam optimizer with a fixed learning rate of 3 × 10⁻⁴ and batch size of 10, and finally early stopping was applied to prevent overfitting. Once the architecture and hyperparameters were completed, the 6-mer count vector from feature engineering was used as the input feature for training the CNN classifier. Upon training, the model learned a set of weights and biases, making it ready for predictions.

**Figure 1.**
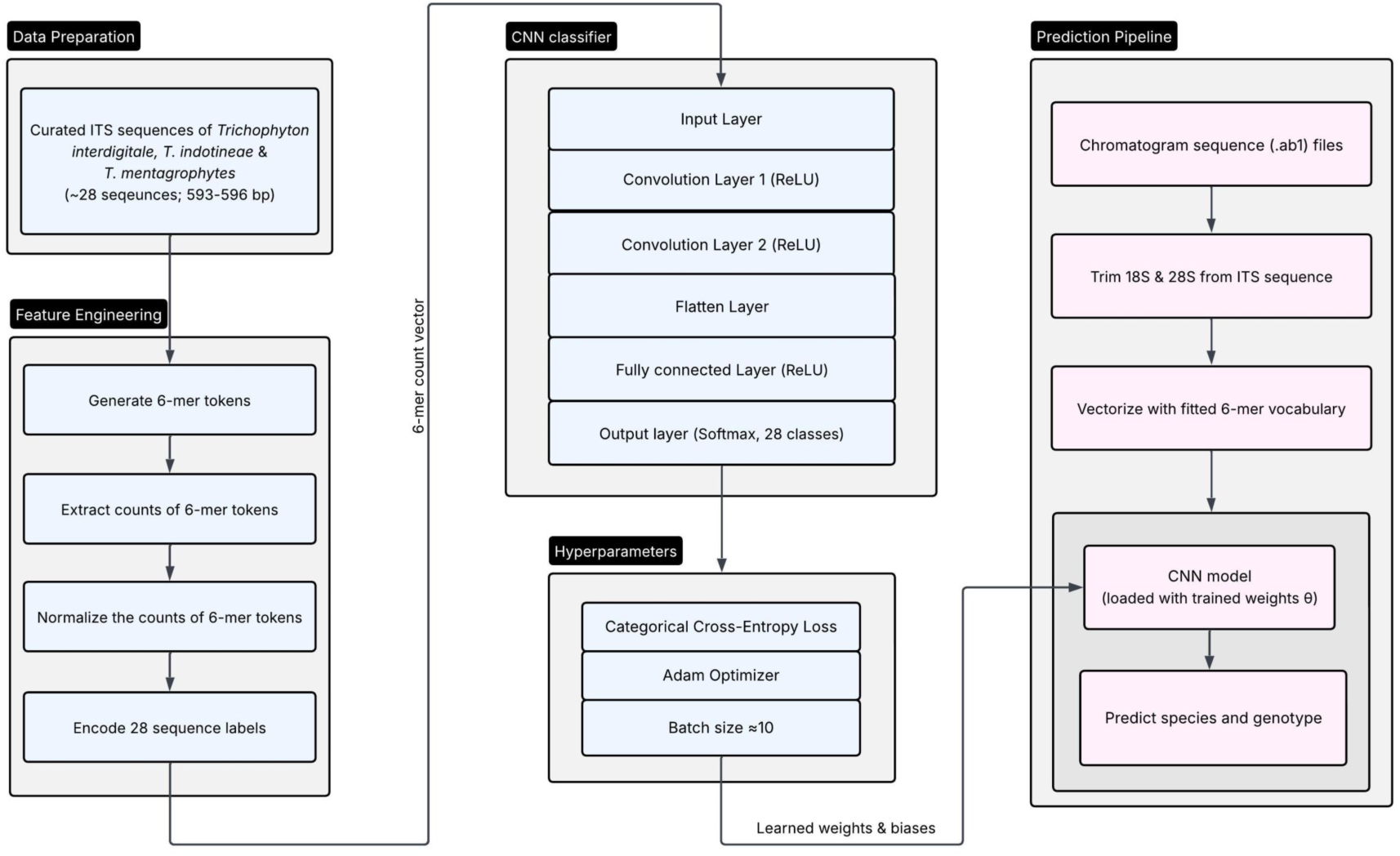
Architecture of the convolutional neural network (CNN) model based ITS sequence pipeline for determination of *TiTm*SC. The diagram illustrates the end-to-end data transformation from raw sequence input to predicted genotype. In brief, the pipeline input nucleotide sequences were first converted into 6-mer token, which passed through two convolutional layers with ReLU activations. Feature maps are pooled and flattened before entering fully connected layers for species and genotype prediction.

Output chromatogram files (.ab1) from Sanger sequencing of the ITS region with forward (V1827) and reverse (V50) primers were directly uploaded into this pipeline. Base calls and per-base phred scores were extracted with Biopython (SeqIO), low quality bases were removed from both the 5’ and 3’ ends using a sliding window approach (window size = 5 bps) with a pred quality score cutol ol ≥ 8. Reverse strands were complemented and aligned to the forward sequence using Python library Pairwise Aligner to generate a consensus sequence. Finally, for the prediction pipeline, each input sequence was subjected to trimming of the conserved 18S and 28S sequences, by searching for conserved motifs at the 5’ ([GA]CGCGCAGGCCGGA[GC]GCTGGCC[GC]CCCACGA) and 3’ (GGCCTCA[AG]AATCTGTTTTATACTTAT[TC][GA]) edges of the ITS region (Supplementary Figure 1). The full-length ITS sequence was vectorized using the fitted 6-mer vocabulary and scaled to generate standardized feature representations. The count vectors were then fed into the CNN model to provide species and genotype identification with a confidence score (0.982 to 1.0) computed from the SoftMax probabilities. To minimize the risk of incorrect genotype assignment, sequences passed to the model that do not belong to any known reference genotype were filtered out using an out-of-distribution detection method (35). For each sequence, we computed the Mahalanobis distance to all classes and only accepted sequences below a threshold of 1. If there is a single SNP from one of the 28 genotypes, the pipeline will provide the most closely related genotype and the SNP position. Any close matches with more than one SNP will not pass the Mahalanobis threshold and give the output “Not a member of the *TiTm*SC, check sequences manually”. If the full-length ITS sequence was not obtainable, the pipeline will give the output “Insulicient sequence for genotyping”. The pipeline code can be found in Supplementary File 1.

### Whole-Genome Sequencing and Bioinformatic Analysis

To determine relatedness among individual *Trichophyton* species and genotypes, 58 *Ti*, 40 *Tm*VII, and 14 *Tm*II* isolates whose identification was confirmed by ITS sequencing were sent for whole genome sequencing at the Wadsworth Center Advanced Genomic Technologies Cluster. Genomic DNA was extracted as described above, libraries were prepared using Illumina NextSeq 500/550 Mid Output Kit v2.5 (300 Cycles, Illumina, Cat# 20024905) and sequenced on either an Illumina NextSeq 500/550 or 1000 instrument (Illumina). Bioinformatic analyses were performed in CLC Genomics Workbench using the CLC Microbial Genomics Module (version 25.0.3; Qiagen, Inc.). Whole genome assemblies were prepared *de novo* for each isolate (Supplementary Table 1-3). All of our *Ti* isolates were mapped to TIMM20114 (GCA_023065905.1), all *Tm*II* isolates were mapped to LL-2024a (GCA_047301425.1), and *TmVII* isolates were mapped to M8436 (GCA_900162065.1) (Supplementary File 2). SNPs were detected and used to construct phylogenetic trees using a maximum likelihood algorithm and a Jukes-Cantor substitution model with 1000 bootstrap replicates. Sequences of SQLE, *CYP51A* and *CYP51B* were extracted for each isolate and variants were identified.

## RESULTS

### ITS Sequencing is Crucial for *Trichophyton* Species and Genotype Determination

Traditional culturing, microscopy, molecular assays, and MALDI-TOF MS have limitations in distinguishing among *Trichophyton* species and genotypes (2). This highlights the practical importance of ITS sequencing in identifying *Trichophyton* species at the genotype level. The entire length of the ITS region (ITS1+5.8S rRNA+ITS2), comprising all unique *TiTm*SC SNPs, was essential for speciation and genotyping. Importantly, the first nucleotide of ITS1 was critical for differentiating *T. interdigitale* (G) from *T. mentagrophytes* (A) (Supplementary Figure 1). Manual alignment of ITS sequences confirmed a total of 151 *Ti,* 42 *Tm*VII, and 14 *Tm*II*\** during the study period from May 2022 to December 2024. Additionally, three isolates of *T. interdigitale* I, 159 isolates of *T. interdigitale* II, one isolate of *Tm* III*, two isolates of *Tm*IV, and one isolate of *T. interdigitale* XI were also identified during this time. Nine isolates matched closely to one of the 28 *TiTm*SC genotypes, but they contained a unique SNP and, therefore, could not be assigned as one of the 28 *TiTm*SC genotypes (data not shown).

### An Automated ITS Analysis Pipeline Enables Rapid and Accurate Identification of *Trichophyton* Species and Genotypes

Given the labor intensive, detailed analyses required to genotype these pathogens via manual alignment of ITS sequences, we developed a pipeline to directly accept Sanger sequencing files as input, trim and align the sequences, and then utilize a deep learning CNN model with 28 curated ITS sequences of the *TiTm*SC (5) to assign a *Trichophyton* genotype. The model was evaluated with 382 clinical isolates of *Trichophyton* received in the laboratory from May 2022 to December 2024. The deep learning CNN model accurately predicted *Trichophyton* species and genotypes with 98.7% concordance to the manual alignment approach, confirming its excellent performance (Figure 2). Nine isolates were closely matched to one of the 28 genotypes but had a single unique SNP difference from the closest reference sequence. Five isolates had underlying background in the chromatogram files (Supplementary Figure 2) and thus did not pass through the pipeline, but were able to be manually inspected and assigned a genotype. In addition, the pipeline also allowed for simultaneous analysis of multiple ITS sequences at once, making the analysis much faster and more efficient.

**Figure 2.**
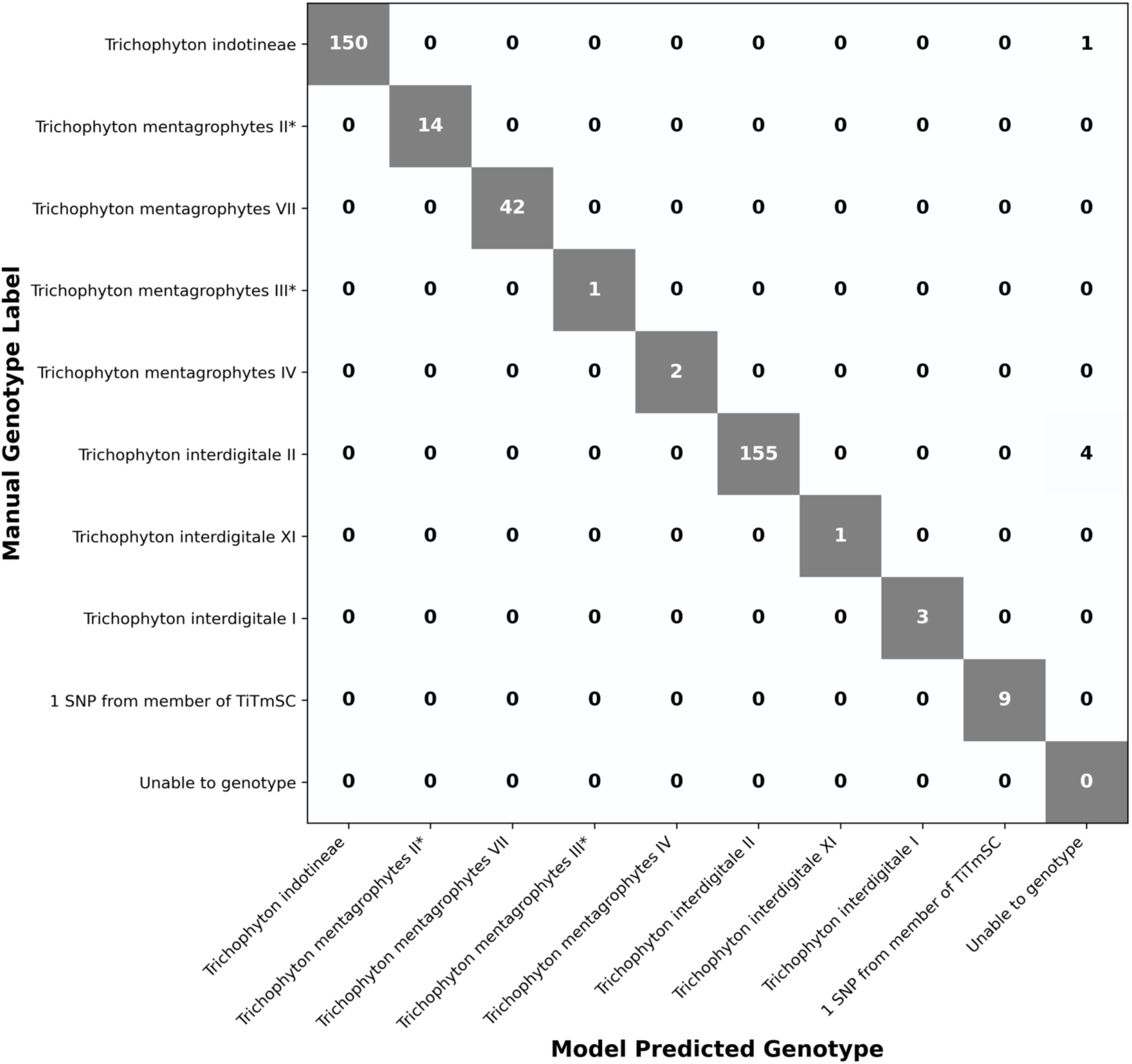
Confusion matrix of the convolutional neural network (CNN) model predicting *TiTm*SC. The vertical axis represents the genotype assigned through manual alignment and the horizontal axis represents the genotypes predicted by the model. There was 98.7% concordance between the two analysis methods. Diagonal entries (shaded boxes) represent isolates correctly identified by the pipeline and off-diagonal entries indicate isolates that could not be automatically genotyped via the pipeline.

### A Rise in *Trichophyton* Cases in New York

Over the past three years, we have seen a dramatic increase in *Trichophyton* cases in NYC and its surrounding counties (Figure 3). We have identified 151 *Ti* isolates representing 135 cases that were concentrated in NYC’s four counties (Kings, Queens, Bronx and New York) while the other nine cases were from the periphery of NYC. The first *Ti* case was identified in May of 2022, followed by one more case in the same year. The number substantially increased to 36 cases in 2023 and 97 cases in 2024 (Figure 4 a). Almost equal numbers of male and female patients were infected with *Ti* (Figure 4 b), and those infected with *Ti* ranged in age from 4 to 79 (Figure 4 c).

**Figure 3.**
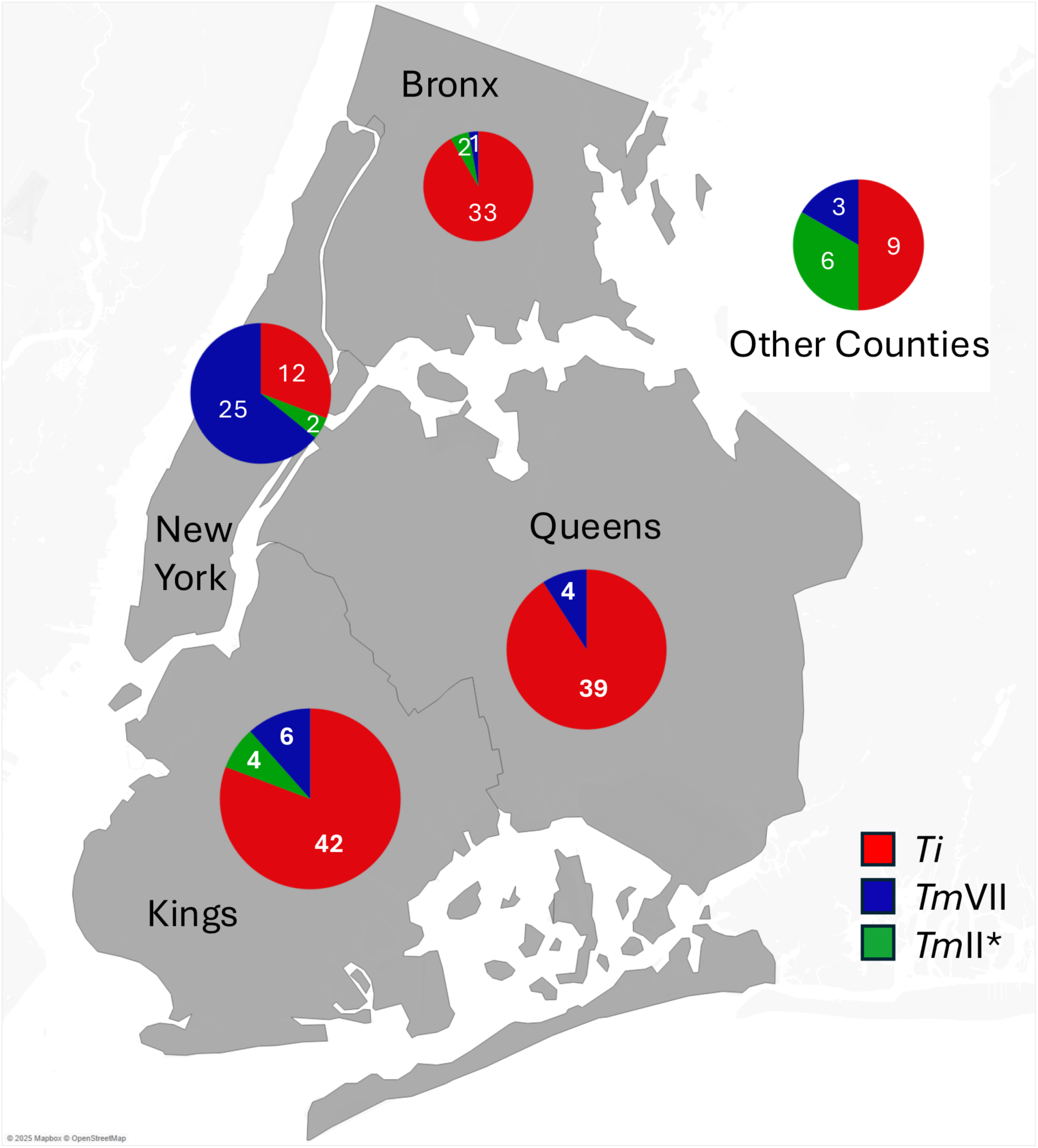
Distribution of *Ti*, *Tm*VII and *Tm*II* cases in NYC and surrounding counties. A map of NYC counties with overall distribution of *Ti* (red), *Tm*VII (blue), and *Tm*II* (green) cases shown as pie chart. The cases clustered mostly in New York, Bronx, Kings and Queens counties. A few additional cases were identified in counties surrounding NYC and are grouped together in the pie chart outside the NYC map.

**Figure 4 a-c.**
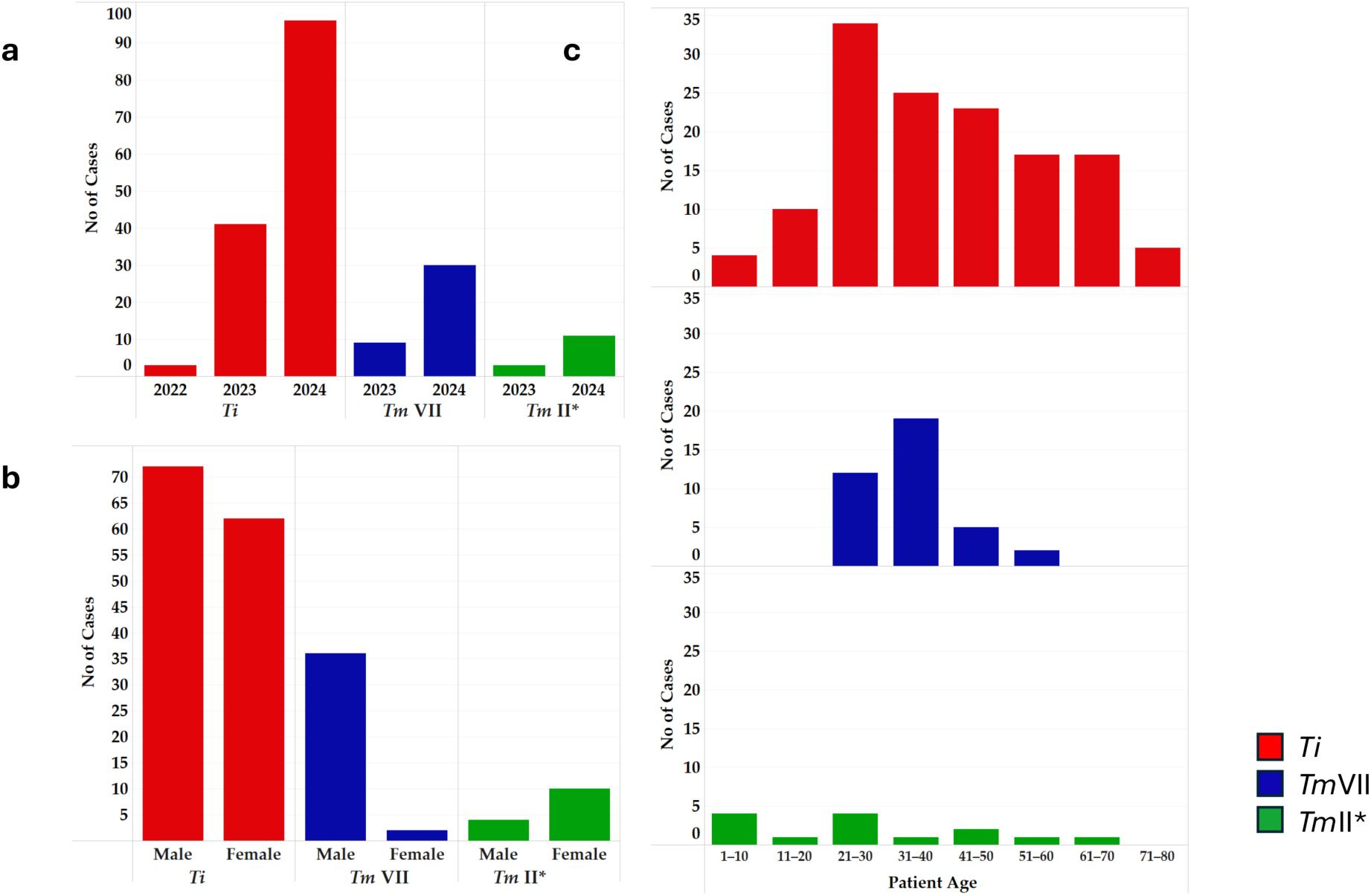
Demographics of *Ti, Tm*VII and *Tm*II* cases in NYC and surrounding counties. a, number of cases identified since 2022. b, sex distribution of cases of *Ti*, *Tm*VII, and *Tm*II*. C, age range for patients with *Ti*, *Tm*VII, and *Tm*II* infections.

During this period, we also identified 42 isolates of *Tm*VII representing 39 cases. Most of the cases were identified from New York County (25 cases) followed by 14 cases from other NYC counties (Figure 3). The first *Tm*VII case was identified in June of 2023 and the total number of cases reached seven by the end of that year. The number of *Tm*VII cases rose substantially in 2024 by 32 cases (Figure 4 a). Patients’ age ranged from 25 to 45 (Figure 4 c), and male patients outnumbered female patients; gender was not specified for one patient (Figure 4 b).

While developing our *TiTm*SC genotyping pipeline, we unexpectedly identified 14 isolates of *Tm*II* representing 14 cases, and these cases were scattered throughout NYC and its surrounding counties (Figure 3). Three *Tm*II*cases were identified in 2023, while the other 11 cases were identified in 2024 (Figure 4 a). Patients’ ages ranged from 5 to 63 (Figure 4 c) and, interestingly, female patients outnumbered male patients (Figure 4 b).

### Emerging Antifungal Resistance in *TiTm*SC

One hundred thirty three isolates of *Ti,* 39 isolates of *Tm*VII, and 10 isolates of *Tm*II* were tested against a panel of antifungal drugs by a broth microdilution assay according to Reference Method M27-A3 of the CLSI guidance document (36). Resistance was determined by available epidemiological cutoff values (ECV) for certain drugs (17, 32, 33). More than half of the *Ti* isolates were resistant to the first line drug, terbinafine (57%), followed by voriconazole (52%), fluconazole (38%), and ketoconazole (31%). Resistance to the second-line drug, itraconazole was minimal (5.3%, Table 1). Furthermore, *Ti* isolates generally fell into three distinct categories of terbinafine resistance: susceptible (≤0.03 μg/mL; 41%), moderately resistant (0.5 – 2 μg/mL; 17%) or highly resistant (≥16 μg/mL; 40%) (Table 1). The Squalene epoxidase encoding SQLE gene was extracted from *de novo* assembled whole genomes of *Ti* and all but one of the moderately resistant isolates contained the mutation L393S, while all highly resistant isolates contained the mutation F397L (Table 1). The SQLE mutations A448T, F415Y, R413G, S443P, F415C were also observed but did not appear to impact terbinafine resistance (data not shown).

To our knowledge, no ECVs have been determined for *Tm*VII or *Tm*II*. If we assume their ECVs are the same as *Ti*, only a small number of *Tm*VII isolates were resistant to voriconazole (5%), fluconazole (13%) or ketoconazole (21%). For *Tm*II*, only one isolate was at the ECV for ketoconazole (10%); all *Tm*II* isolates were susceptible to all other drugs. Importantly, neither *Tm*VII nor *Tm*II* exhibited any resistance to the first-line drug terbinafine or second-line drug itraconazole.

### WGS Revealed Clonal and Non-Clonal Spread of *Trichophyton* Species and Genotypes

To understand the spread of *Ti*, *Tm*VII, and *Tm*II* in patients living in NYC and its periphery, we performed WGS on a select number of isolates (58, 40 and 14, respectively). Genomes were assembled *de novo* and assembly characteristics are presented in Supplementary Tables 1-3. The average genome size for *Ti*, *Tm*VII, and *Tm*II* isolates was 22.13, 22.58 and 22.63 Mbp, respectively. The details of how a reference strain was chosen for *Ti*, *Tm*VII and *Tm*II* are provided in Supplementary File 2.

Mapping of 55 *Ti* isolate reads to the reference TIMM20114 isolate resulted in four major clusters with SNPs ranging from 0-415 and the SNP difference within each cluster was <108 (Figure 5). Also, within each cluster, we found a few pairs of isolates with 0-3 SNPs (Figure 5), suggesting a possible clonal spread. In the absence of epidemiological data for majority of *Ti* isolates that were sequenced, we investigated whether patient geographic proximity played any role in the spread of *Ti* among patients. We evaluated 17 isolates originating from 17 patients living within a single zip code. The results revealed that *Ti* isolates grouped into three distinct clusters (Supplementary Figure 4), with SNP differences ranging from three to 138. These results suggested that, except in a few instances of clonal spread, most cases were due to non-clonal spread of *Ti*.

**Figure 5:**
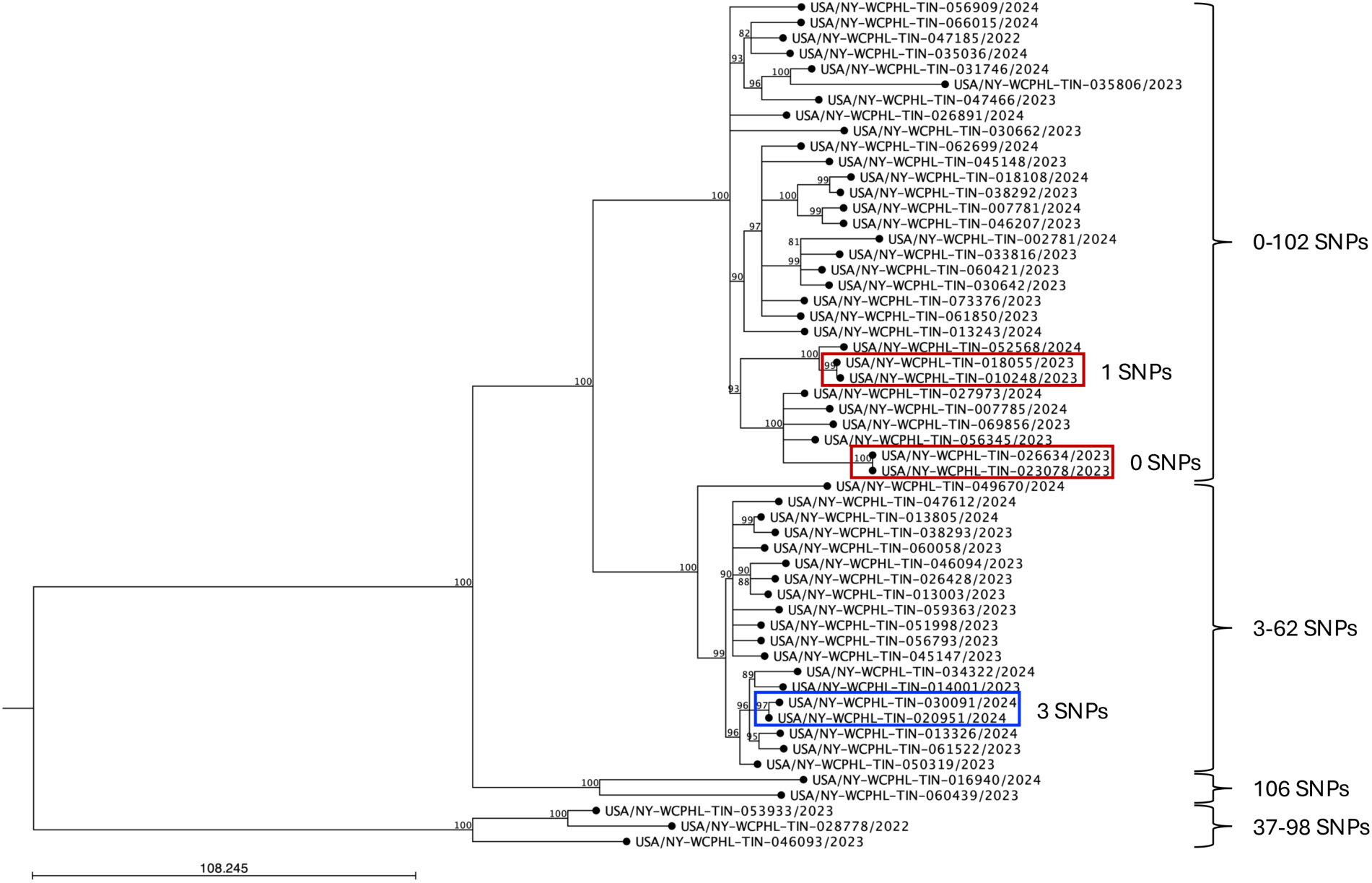
Maximum likelihood SNP tree of *Ti* isolates. *Ti* reads were mapped to the reference strain TIMM20114, and a phylogenetic tree was constructed using a maximum likelihood algorithm with a Jukes-Cantor substitution model and 1000 bootstrap replicates. A bootstrap cutoff of 80% was used and bootstrap values are indicated at nodes. The scale bar represents number of expected substitutions between samples. All *Ti* isolates grouped into four clusters. SNP ranges in each cluster are provided next to the brackets on the right. Confirmed cases of household transmission are boxed in red from previous investigation (15) and a third instance of likely local transmission is boxed in blue in current investigation.

The *Tm*VII isolates from the current study were mapped to the GenBank assembly strain M8436, which was confirmed as *Tm*VII by our pipeline. Contrary to our results for *Ti*, all but two isolates of *Tm*VII grouped in the phylogenetic tree with <19 SNP differences among them, indicating possible clonal spread of *Tm*VII in the greater NYC area (Figure 6). One isolate fell outside this main cluster and was 16-27 SNPs from isolates in the main cluster. The second isolate was from a patient who resided in a county with no other cases and was quite distinct from the rest of the *Tm*VII isolates, with SNP differences ranging from 123 to 135 (Figure 6).

**Figure 6.**
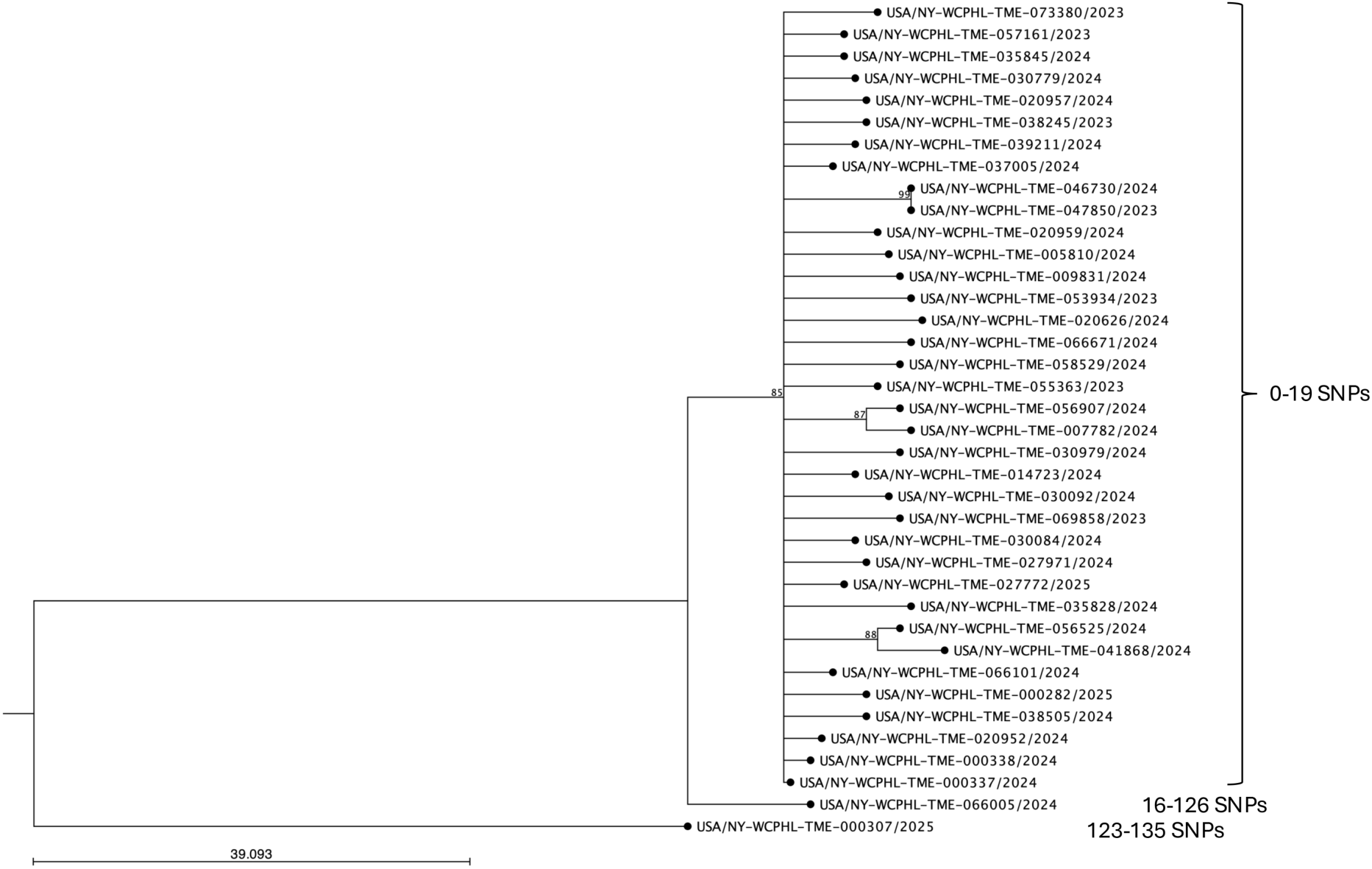
Maximum likelihood SNP tree of *Tm*VII isolates. *Tm*VII reads were mapped to the reference strain M8436 and a phylogenetic tree was constructed using a maximum likelihood algorithm with a Jukes-Cantor substitution model and 1000 bootstrap replicates. A bootstrap cutoff of 80% was used and bootstrap values are indicated at nodes. The scale bar represents the number of expected substitutions between samples. All but two isolates of T*m*VII grouped into one cluster with <19 SNP differences among them, indicating possible clonal spread in the greater NYC area. One isolate fell outside the main cluster with 16-27 SNPs apart from the main cluster. The second isolate was from a patient who resided in a county with no other cases and was quite distinct from the rest of the *Tm*VII isolates, with SNP differences ranging from 123 to 135. SNP ranges in each cluster are provided next to the brackets on the right.

LL-2024a was selected as the reference strain for *Tm*II* read mapping. The resulting SNP tree (Figure 7) revealed one cluster of seven isolates that contained two instances of likely clonal transmission (zero and three SNPs between isolate pairs), and overall, the cluster contained ≤47 SNPs. Other isolates of *Tm*II* had 99-687 SNPs in common with all other isolates and did not appear to be related to any other isolates in this cohort. A *Tm*II* isolate with a single SNP (G220A) was also sequenced and found to be 113-558 SNPs from otherTmII* isolates.

**Figure 7.**
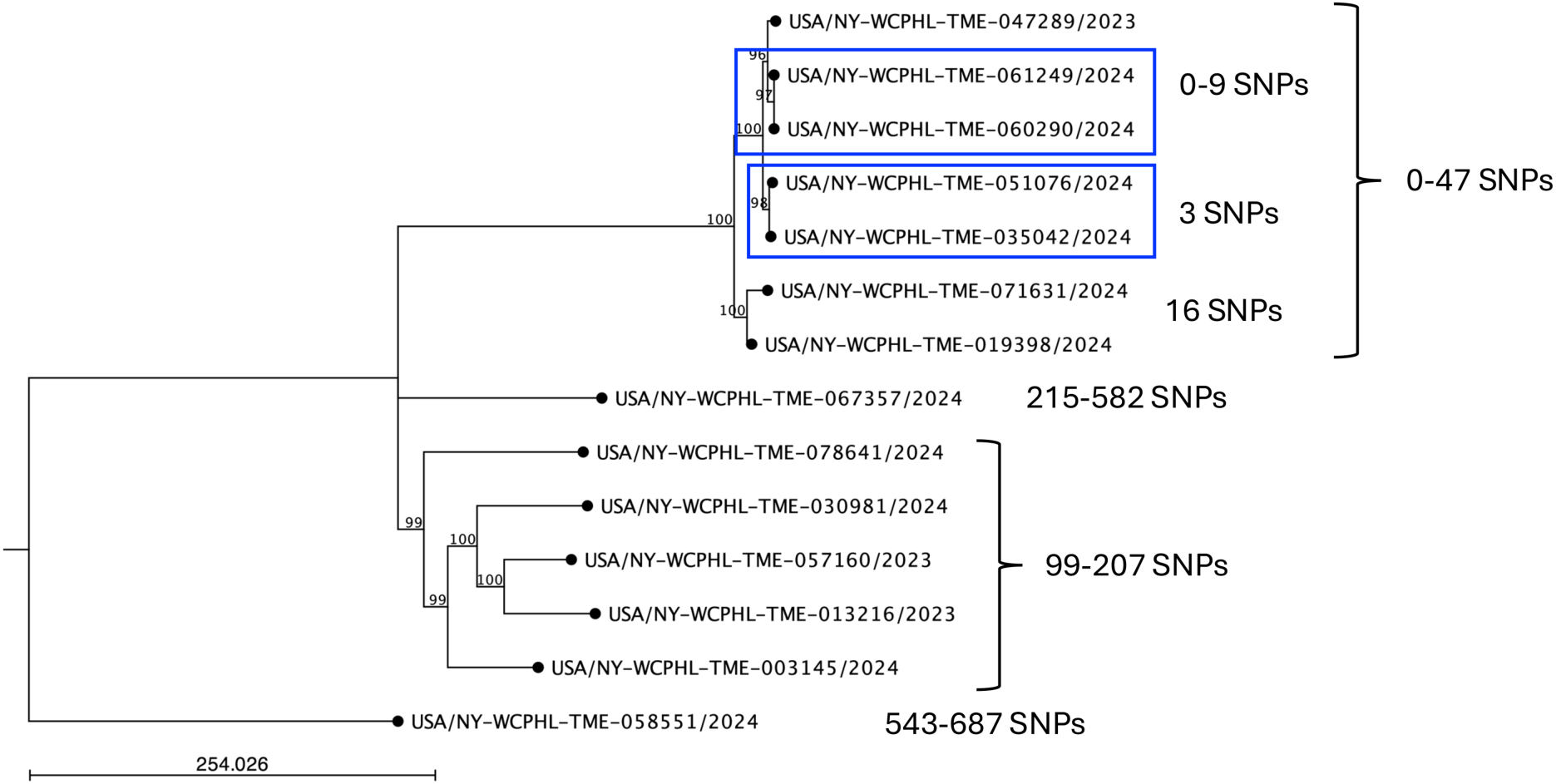
Maximum likelihood SNP tree of *Tm*II* isolates. *Tm*II* reads were mapped to the reference strain LL-2024a, and a phylogenetic tree was constructed using a maximum likelihood algorithm with a Jukes-Cantor substitution model and 1000 bootstrap replicates. A bootstrap cutoff of 80% was used and bootstrap values are indicated at nodes. The scale bar represents the number of expected substitutions between samples. The *Tm*II* isolates clustered into four distinct group. SNP ranges in each cluster are provided next to the brackets on the right. Blue boxes indicate potential cases of local transmission.

## DISCUSSION

This report presents a comprehensive laboratory investigation of emerging drug-resistant *Ti*, *Tm*VII, and *Tm* II* from NYC and surrounding counties. These findings have significant implications for infection control, providing valuable insights into the spread of these pathogens and guiding future infection control measures. We previously documented our early cases of *Ti* (14, 15), which mainly involved patients who had recently traveled to, or had family members who had traveled to, Bangladesh. Since our last report, there has been a rapid rise in *Ti* cases, which is a concerning trend. It is also important to mention here that *Ti* does not appear to be gender or age-specific, as both male and female patients, ranging in age from 4 to 76, were impacted by this pathogen. Our WGS analysis on a random selection of *Ti* isolates revealed four major clusters. However, relatedness among *Ti* isolates in each cluster cannot be assessed, as there is currently no SNP cutoff established to delineate relatedness for dermatophytes. When sequencing multiple isolates from the same patient, we and others have observed only zero to two SNPs between isolates (16). Based on our previous work, we know of two cases of household transmission where there were zero to two SNPs between patients’ isolates (Figrue 5, red boxes); other isolates from that study were from patients who had recently travelled to, or had a family member who travelled to, Bangladesh and had 32-373 SNPs from other *Ti* isolates, which we interpreted to be travel-related, independent introductions (15). In the current study, we identified an additional pair of NYC *Ti* isolates that differ by only three SNPs (Figure 5, blue box), which likely represents a local transmission event. However, it is unclear whether the higher SNP values between *Ti* isolates in each of the four major clusters represent local transmission or independent introductions related to travel. A study of *Ti* isolates from five hospitals in North India over a five-year period found only 42 SNPs among all isolates, concluding that these were likely caused by clonal spread (37). A group in Singapore identified two *Ti* isolates with 92 SNPs between them and <182 SNPs from other isolates collected from patients in India (38). Finally, a global analysis of *Ti* isolates found that the first 11 *Ti* isolates identified in NYC were dispersed throughout multiple clusters of *Ti* from Canada and around the world with 0-375 SNPs (15, 16). The lack of geographic clustering strongly suggests that, aside from a few cases of local transmission, most of our cases are likely independent introductions resulting from global travel.

Following our initial report of *Tm*VII infections (28), we have also observed a rapid increase in cases of *Tm*VII in NYC. In contrast to *Ti* infections, the rise in *Tm*VII cases appears to be primarily due to clonal spread, as most isolates were clustered in the phylogenetic tree with less than 19 SNPs apart. Although *Tm*VII infections were predominantly impacting men, we did find a few cases of women infected with this pathogen. The precise mechanisms leading to the spread of *Tm*VII infection are currently unclear; infections appear to be primarily spreading through direct skin-to-skin contact during sexual activity, pubic hair grooming, or regular use of shared spaces like gyms and saunas (1, 25). These findings underscore the need for further research to understand the dynamics of *Tm*VII infection and spread.

In this investigation, we have also identified 14 patients infected with *Tm*II*, which appears to be the first report of this pathogen from NYC as well as from the US. Patients either had tinea capitis (scalp infection) or tinea corporis (ringworm on the body). The unique ability of *Tm*II* to infect the scalp or body, a characteristic that set it apart within the taxonomically complex *TiTm*SC was observed (1). Another uniqueness observed was that the female patients outnumbered male patients with *Tm*II* infection, which is consistent with a single published report, although that report consisted of only two cases of *Tm*II* (1). These results emphasize the need for clinicians to remain vigilant regarding evolving genotypes, atypical clinical presentations, and antifungal resistance to ensure timely diagnosis and appropriate treatment for *Trichophyton* infection.

Clinical breakpoints for antifungal drugs are currently not available for any dermatophyte. However, we have observed that 57% of our NYC *Ti* isolates had a moderate to high MIC to the first-line drug, terbinafine, which correlated with the mutations L393S and F397L in SQLE, respectively. These findings are not surprising, as several studies have reported that *Ti* is increasingly found to be resistant to terbinafine due to mutations in SQLE at positions L393, F397, F415, and H440 (1, 13, 15, 18, 20, 32, 37, 39–41). Interestingly, however, isolates resistant to terbinafine did not cluster in the *Ti* phylogenetic tree (data not shown), indicating that resistance likely develops independently due to exposure to or inappropriate use of antifungals (41). Decreased susceptibility to azoles was also observed in some *Ti* isolates, raising concerns over the potential evolution of multidrug-resistant isolates, which would further complicate patient care. Azole resistance has been associated with the overexpression of or mutation in the *CYP51B* gene, which encodes a sterol 14α-demethylase (42). While some SNPs in Cyp51B were found, there was no correlation between any SNP(s) and elevated azole MICs (data not shown), which is similar to results reported by others (16, 19). However, we cannot rule out the possibility of Cyp51B overexpression due to gene multiplication, a previously documented resistance mechanism identified via long-read sequencing (16, 42). Other mechanisms of resistance are currently unknown, but overexpression of transporters, other drug efflux pumps, biofilm formations, and loss of mitochondrial complex have been linked to azole resistance (43, 44).

To date, we have observed no resistance to terbinafine in our *TmVII* or *Tm*II* isolates and only a low level of resistance to azoles. However, given the overuse of topical combination creams and empirical treatment of infections prior to organism identification and AFST, it is a matter of time before these pathogens develop resistance. Therefore, AFST should be made easily accessible across various healthcare settings to ensure the appropriate selection of antifungal drugs, thereby reducing treatment duration and patient suffering.

Although there are apparent differences in infection characteristics and antifungal resistance profiles among *TiTm*SC members, it remains challenging to identify isolates to the species and genotype level. Molecular techniques provide a high degree of accuracy in species and genotype identification, though their use can be limited by cost and availability. A real-time PCR assay for the identification and detection of *Ti* from isolates and primary specimens was recently reported (45), which will aid in early diagnosis and treatment for patients with suspected *Ti* infection. In addition, a comparative study of various dermatophyte diagnostic tests with several members of the *TiTm*SC revealed that while most techniques were able to identify an isolate as *Trichophyton* spp., none could distinguish among *TiTm*SC genotypes except MALDI-TOF MS identification of *Ti* (2, 5). However, while some groups have achieved a 96% identification rate for *Ti* by MALDI-TOF MS, there is currently no commercially available MALDI database containing *Ti* spectra or other members of the *TiTm*SC at the genotype level.

Currently, ITS sequencing is regarded as the gold standard for dermatophyte identification due to its availability, specificity, and relatively faster turnaround time (15, 28). However, manually analyzing *TiTm*SC ITS sequences and correctly assigning species and genotype can be time-consuming and tricky, especially since high-quality sequence spanning the entire ITS region is required for accurate speciation and genotyping (Supplementary Figure 1). While bioinformatic pipelines exist to aid in genotype identification (6), they do not remove the cumbersome task of manually trimming ITS sequences and they cannot provide identification at the genotype level if sufficient sequence in not available. In this work, we created a completely automated pipeline that uses the output chromatogram files from Sanger sequencing as inputs and will automatically clean/trim sequences, extract a consensus sequence of sufficient length (if sequence quality is high enough), and match it to one of the 28 genotypes in the *TiTm*SC. This pipeline eliminates the need for manual sequence processing, thereby expediting dermatophyte identification, particularly during large outbreaks.

Given the complexity of distinguishing among *TiTm*SC members, it is unsurprising that none of the genome assemblies in GenBank have been identified to the genotype level, except *Ti*, and many described as *T. interdigitale* or *T. mentagrophytes* are speciated incorrectly. We have utilized our pipeline to accurately assign species and genotypes to the *T. interdigitale*, *T. mentagrophytes*, and *Ti* genome assemblies in GenBank (Supplementary Table 4), making this information accessible to the broader scientific community and drawing attention to the distinct infection characteristics among *TiTm*SC members.

During our retrospective analysis of all 382 *TiTm*SC isolates, we found nine that did not have a 100% match (one SNP) to any of the currently designated *TiTmS*C ITS genotypes. Whole-genome sequencing to determine ANI and relatedness to other members of the *TiTm*SC, along with unique clinical presentations or resistance patterns, will be critical before usefully demarcating more genotypes in the *TiTmS*C.

In conclusion, the rapid spread of drug-resistant *Ti*, sexually transmitted *Tm*VII, and newly emerging *Tm*II* underscores the need for accurate species and genotype-level identification. Additionally, increasing reports of antifungal resistance and treatment failure necessitate strengthening antifungal stewardship, particularly when using systemic antifungals in combination with or without topical antifungals, or with or without steroid creams. We have observed both clonal and non-clonal transmission of these pathogens, emphasizing the ease with which these pathogens spread. A better understanding of *Ti, Tm*VII*, and Tm*II*\** transmission pathways is needed as well as education for clinicians, other healthcare professionals, and patients. The comprehensive management of these infections should include the right choice of antifungals, use of AFST for determining *in-vitro* resistance, elimination of environmental reservoirs, clear clinical guidelines, and patient education on household hygiene and effective textile disinfection strategies to prevent transmission, re-infection, and unnecessary antifungal use.

## Data Availability

All data produced in the present work are contained in the manuscript

---

**Supplementary Figure 1. Alignment of ITS sequences for the 28 genotypes of the *TiTm*SC.** The 5.8S ribosomal sequence is in red, internal transcribed spacers (ITS) 1 and 2 are in blue, and primer binding sites in conserved regions of the 18S and 28S rRNA for forward (V1827) and reverse (V50) primers, respectively, are in green. SNPs among the 28 genotypes are highlighted in pink.

**Supplementary Figure 2.** Two examples of background in the Sanger sequencing chromatogram. In these situations, manual inspection is required for accurate genotyping.

**Supplementary Figure 3. Determining the best reference strain.** (a) K-mer analysis of a representative from our *TiTm*SC isolate assemblies along with seven GenBank assemblies. A *T. rubrum* isolate is included as an outgroup. (b) Matrix of the average nucleotide identity (ANI) between between GenBank assemblies and representatives from our *TiTm*SC isolate assemblies. A *T. rubrum* isolate is included as an outgroup.

**Supplementary Figure 4. Maximum likelihood SNP tree of Ti isolates from patients living in a single zip code.** Ti reads were mapped to the reference strain TIMM20114 and a phylogenetic tree was constructed using a maximum likelihood algorithm with a Jukes-Cantor substitution model and 1000 bootstrap replicates. A bootstrap cutoff of 80% was used and bootstrap values are indicated at nodes. The scale bar represents the number of expected substitutions between samples. SNP ranges in each cluster are provided next to the brackets on the right, and an instance of likely local transmission is boxed in blue and labeled with the SNP differences between those isolates.

## Authors Contributions

GT contributed to the AFST and WGS testing, data analysis, writing and editing of the manuscript, and prepared graphs and tables. VV developed and optimized a bioinformatics pipeline with ITS sequences, wrote a section of the manuscript, and prepared figures. BO performed the ITS-PCR/sequencing, manual sequence alignment, identification, and data analysis. SC contributed to the conception and design of the study and wrote the first and final draft of the manuscript. All authors contributed to the manuscript and approved the submitted version.

## Acknowledgements

We thank Wadsworth Center Media and Tissue Culture Core for providing all media necessary for culturing of dermatophytes. We thank Wadsworth Center Advanced Genomic Technologies Core for dermatophyte DNA sequencing on Sanger and Illumina platforms. Our very special thanks to Drs. Spencer Bruce and Jaida Li for their careful review of bioinformatics pipeline. We also want to thank Dr. Lisa A Biega for critical reading of the manuscript. This study was partially supported by the Centers for Disease Control and Prevention Epidemiology and Laboratory Capacity Antibiotic Resistance Laboratory Network Grant (NU51CK000372) and CDC’s Advanced Molecular Detection Grant (GR150201607).

